# The density of routinely collected neurology data depends on patient visit type: an investigation using the Observational Medical Outcomes Partnership Common Data Model

**DOI:** 10.1101/2025.05.12.25327416

**Authors:** Fran Biggin, Laura White, Quin Ashcroft, Tim Howcroft, Vishnu Chandrabalan, Hedley Emsley, Jo Knight

**Affiliations:** Lancashire Teaching Hospitals NHS Foundation Trust; Lancaster University

## Abstract

**Background:** The Observational Medical Outcomes Partnership (OMOP) Common Data Model (CDM) is a standardised framework for organising healthcare data. This study investigates the practicality of using the OMOP CDM to analyse data on neurology patients.

**Methods:** An outpatient neurology patient cohort was defined on the basis of having attended at least one neurology outpatient appointment between 01 April 2022 and 31 March 2023 (n=23,862). All data collected at visits made by this cohort between 01 April 2021 to 31 March 2024 was extracted. The cohort was then divided into 4 sub-cohorts according to appointment types attended: outpatient appointment(s) only (n=15,255); outpatient appointment(s) and inpatient stay(s) (n=2750); outpatient appointment(s) and emergency department attendance(s) (n=1658); outpatient appointment(s), inpatient stay(s) and emergency department attendance(s) (n=4199).

**Results:** We found there to be more OMOP-mapped data available for patients who had at least one inpatient stay or emergency department attendance than for those with only outpatient appointments. Notably, an average of 0 out of 100 patients in the outpatient only sub-cohort had a record of a condition, compared to 100 out of 100 patients in the sub-cohort with outpatient appointments, emergency attendances and inpatient stays.

**Conclusions:** Neurology outpatients have far less data recorded than inpatients or patients attending ED. This disparity arises from the lack of outpatient diagnostic coding and impairs the advancement of research in this area.

**KEY MESSAGES:** *What is already known on this topic:* the OMOP common data model (CDM) is being adopted by the NHS to provide a uniform structure to the data within the NHS Secure Data Environments to support research. We know that outpatient coding is not mandated so diagnoses at outpatient appointments are not regularly recorded in EHRs.

*What this study adds:* we investigate the variable volume of data available for research through a secondary care dataset that has been converted to the OMOP CDM. We show that outpatients have far less data recorded than inpatients or patients attending ED, in terms of both volume and type of data.

*How this study might affect research, practice or policy:* this study highlights the need for data systems such as SDEs to be based on data which is complete. We also highlight the importance of ensuring that data recording for outpatients is as complete as it is for inpatients and ED.

## INTRODUCTION

Healthcare data is large and is collected for many different reasons ranging from administration and financing to diagnosis and treatment. The structure of the NHS is complex with different regions commissioning services independently, which has created a situation in which there are a multitude of different electronic systems and databases in use both across and within Trusts. This creates a fractured data landscape where data in one hospital is not compatible with data from another, leading to problems when trying to manage patients who have used services from different Trusts, and when trying to amalgamate data for research purposes. One potential approach to solving this problem is the use of Common Data Models (CDMs) which create standardised structures for representing and storing data, allowing for common understanding of concepts and the ability to share data and processes across institutions that use the same model.

The Observational Medical Outcomes Partnership Common Data Model (OMOP CDM) is a data model designed to be used for observational health data. It provides a framework for transforming data from electronic heath records and other sources into a set of standard vocabularies stored in a standardised database structure [1]. This standardisation allows for the creation of multi-partner studies across geographical boundaries as institutions can share analysis pipelines instead of data [2]. It is currently used by a number of institutions across the USA, Europe and Asia [3], and the OHDSI collaborative, who maintain the OMOP CDM standards, have 4,294 collaborators across 83 countries [4]. The OMOP CDM has been chosen by the NHS to form the structure of their network of Secure Data Environments (SDEs) [5].

The North West SDE covers the NHS Integrated Care Systems of Cheshire and Merseyside, Greater Manchester, and Lancashire and South Cumbria (L&SC). Lancashire Teaching Hospitals NHS Foundation Trust (LTHTR) which is part of L&SC ICS have been instrumental in creating infrastructure and transforming data for the North West SDE [6]. Several of the Trust’s databases containing patient data have been transformed into the OMOP CDM format. In this study we focus on the Trust’s data for neurology outpatients that has been transformed into the OMOP CDM.

Neurology data has been a focus of interest for LTHTR for the past 5 years and previous studies have shown that neurology outpatients in particular lack data [7]. Since many neurological diseases are chronic in nature [8], a high proportion of patients require long-term monitoring in the outpatient setting. The paucity of diagnostic coding for neurology patients who are seen solely in outpatient clinics creates a barrier for clinical research and impairs effective hospital resource planning for outpatient services. A lack of outpatient diagnostic coding has also been identified for neurology patients in other UK Trusts [9], and for outpatients in other medical specialties [10,11], in particular paediatrics [12].

In this study we aim to quantify the volume and type of data available for neurology outpatient appointments, especially when compared to other types of visit (inpatient stays and emergency department attendances). It is important to understand what type of data is currently available as this can tell us what research is currently possible, and what data may need to be brought into the OMOP CDM database to facilitate future studies and potential clinical uses.

## METHODOLOGY

Between March and October 2023 several databases in Lancashire Teaching Hospitals NHS Foundation Trust (LTHFT) were transformed into the OMOP CDM format through a detailed extract-transform-load (ETL) process [13]. The data were chosen to be transformed based on frequency of use, and the majority of the data in the main Electronic Health Record (EHR) were uploaded to the CDM. In addition, data from the most used fields of the Somerset Cancer Registry and the Swisslab pathology database were also taken into the OMOP CDM. This research makes use of a pseudonymised instance of the LTHFT OMOP database IDRIL-1 [14]. The data used in this study were extracted in April 2024.

All relational databases are structured around the use of tables, with common fields in those tables allowing them to be linked to each other. The OMOP CDM is made up of several standardised tables based on categories of data [15], and uses several fields such as ‘person_id’ or ‘concept_id’ which enable the tables to be linked. The ‘visit occurrence’ table contains administrative information on visits such as when visits occurred, the type of visit, and the location of the visit. The tables which contain the majority of the clinical information are ‘condition occurrence’, ‘drug exposure’, ‘procedure occurrence’, ‘measurement’ and ‘observation’. A description of the OMOP tables used in this study can be found in table 1; descriptions are taken from the OHDSI website [15].

**Table 1:** A description of the OMOP tables used in this study.

| OMOP table name / Data category name as used in this study | Description | Example information contained in table |
| --- | --- | --- |
| condition_occurrence / Condition | This table contains records suggesting the presence of a disease or medical condition stated as a diagnosis, a sign, or a symptom, which is either observed by a provider or reported by the patient. | A diagnosis of epilepsy. |
| drug_exposure / Drug | This table captures records about the exposure to a drug ingested or otherwise introduced into the body | A prescription for levetiracetam. |
| procedure_occurrence / Procedure | This table contains records of activities or processes ordered by, or carried out by, a healthcare provider on the patient with a diagnostic or therapeutic purpose. | A record of an MRI brain. |
| Measurement / Measurement | This table contains records of measurements, i.e. structured values (numerical or categorical) obtained through systematic and standardized examination or testing of a person or person's sample. | The results of a complete blood count. |
| Observation / Observation | This table captures clinical facts about a person obtained in the context of examination, questioning or a procedure. | A record that the patient has a family history of stroke. |
| visit_occurrence / Visit | This table contains events where persons engage with the healthcare system for a duration of time. | A record of an outpatient appointment with a consultant neurologist. |

We identified a cohort of neurology outpatients using the ‘visit occurrence’ table. We searched for all patients with an outpatient clinic appointment between 01/04/22 and 31/03/23 where the ‘visit source value’ was ‘OP’ (outpatient), and the ‘provider id’ contained an id from a list of known neurology specialists. We then extracted all data recorded for these patients between 01/04/21 and 31/03/24 (one year preceding the start of, and one year following the end of, the cohort identification period) from the ‘condition occurrence’, ‘drug exposure’, ‘procedure occurrence’, ‘measurement’ and ‘observation’ tables. Note that all patients have had at least one neurology outpatient appointment during the study period, but that data on visits for **all** reasons were extracted for analysis. The overall cohort is one of neurology outpatients, but we include in the analysis hospital visits for any and all reasons, not just neurology.

We created four sub-cohorts from the initial cohort of neurology outpatients:

1. Outpatient appointments only (OP Only) - patients that have **only** had outpatient appointments and **no** other visit types during the study period 01/04/21 to 31/03/24.
2. Outpatient and Inpatient (OP & IP) – patients that have had at least one outpatient appointment **and** at least one inpatient stay but **no** emergency department attendances during the study period 01/04/21 to 31/03/24.
3. Outpatient and ED (OP & ED) – patients who have had at least one outpatient appointment and at least one emergency department attendance but **no** inpatient stay during the study period 01/04/21 to 31/03/24.
4. Outpatient, inpatient and emergency (OP & IP & ED)– patients who have had at least one neurology outpatient appointment **and** at least one inpatient stay **and** at least one emergency department attendance visit during the study period 01/04/21 to 31/03/24.

We used these sub-cohorts to examine the differences in the volume and type of information available on different groups of patients. We examined the overall volume of data available of different data categories, the median number of records created during a visit-day, and the percentage of patients who could be expected to have at least one record in a particular category of data.

In order to compare the number of records per visit we use the concept of a ‘visit-day’ to ensure that we are accounting for visits that occur over multiple days. A ‘visit-day’ is one day of a visit, so an outpatient appointment that occurs on one day accounts for one visit-day, and an inpatient stay with one overnight occurs over two visit-days. Note that many inpatient stays are day cases and are therefore only account for one visit-day (eg for dialysis or chemotherapy).

## RESULTS

We found 23,862 patients who had at least one neurology outpatient appointment between 01/04/22 and 31/03/23. Between 01/04/21 and 31/03/24 these patients had a total of 262,592 hospital visits, split between outpatient appointments (205,180), inpatient stays (36,631) and emergency department attendances (20,781). These patients are split into four sub-cohorts for further analysis depending on the types of visit they made during the study period:

1. Outpatient appointments only (15,244)
2. Outpatient appointments and inpatient stays (2,750)
3. Outpatient appointments and ED attendances (1,658)
4. Outpatient appointments, inpatient stays and ED attendances (4,199)

We can see that the largest sub-cohort is the group of patients who only visited the hospital for an outpatient appointment, with the 15,244 patients in this sub-cohort accounting for 64% of the total study population.

Table 2 shows the number of individual records present in each of the 5 data categories for each of the four sub-cohorts. We can see from this table that sub-cohort 4 (OP&IP&ED) accounts for 80.7 percent of all the records we retrieved despite it representing only 18% of the study population (over 11 million records for 4,199 patients). Within this sub-cohort, the drugs and measurement categories are the two with the highest percentage representation (85.7% and 82.7% of all records). In contrast sub-cohort 1 (OP Only) represents only 1.3% of the total number of records, despite being the largest sub-cohort.

**Table 2:**
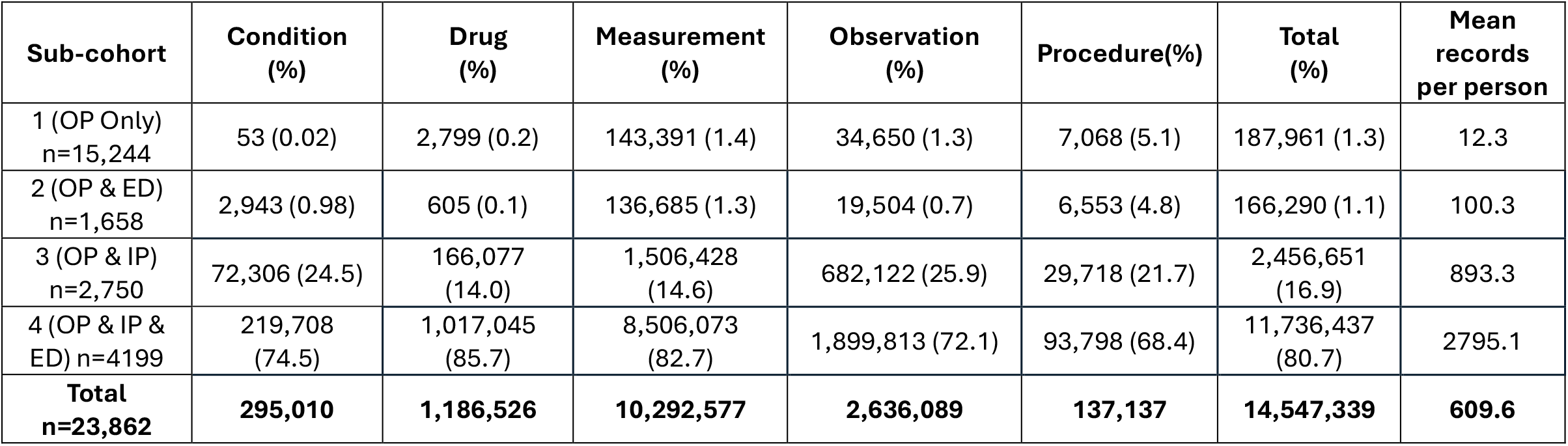
Number of records per sub-cohort in each of the OMOP tables. Note: percentages relate to the total numbers of records in each category, i.e. the column.

| Sub-cohort | Condition (%) | Drug (%) | Measurement (%) | Observation (%) | Procedure(%) | Total (%) | Mean records per person |
| --- | --- | --- | --- | --- | --- | --- | --- |
| 1 (OP Only)<br>n=15,244 | 53 (0.02) | 2,799 (0.2) | 143,391 (1.4) | 34,650 (1.3) | 7,068 (5.1) | 187,961 (1.3) | 12.3 |
| 2 (OP & ED)<br>n=1,658 | 2,943 (0.98) | 605 (0.1) | 136,685 (1.3) | 19,504 (0.7) | 6,553 (4.8) | 166,290 (1.1) | 100.3 |
| 3 (OP & IP)<br>n=2,750 | 72,306 (24.5) | 166,077 (14.0) | 1,506,428 (14.6) | 682,122 (25.9) | 29,718 (21.7) | 2,456,651 (16.9) | 893.3 |
| 4 (OP & IP & ED) n=4199 | 219,708 (74.5) | 1,017,045 (85.7) | 8,506,073 (82.7) | 1,899,813 (72.1) | 93,798 (68.4) | 11,736,437 (80.7) | 2795.1 |
| **Total**<br>**n=23,862** | **295,010** | **1,186,526** | **10,292,577** | **2,636,089** | **137,137** | **14,547,339** | **609.6** |

Table 3 shows the number of hospital visits made by patients in each of the four sub-cohorts. We can see that patients in sub-cohort 4 (OP&IP&ED) had the highest number of all types of visit, including more outpatient appointments than sub-cohort 1 (OP Only) despite the sub-cohort being a third of the size. This may be due to the fact that patients who require both inpatient stays and ED attendances may have more complex conditions and multiple comorbidities, leading to the need for a higher number of outpatient appointments. All patients who had ED attendances that led to inpatient stays are included in sub-cohort 4. There are 5,520 of these types of visit and, for the purpose of this analysis, they are counted twice, both as an ED attendance **and** as an inpatient stay.

**Table 3:** Number of visits attended.

| Sub-cohort | Outpatient Appointments | Inpatient Stays | Emergency Attendances | Total |
| --- | --- | --- | --- | --- |
| 1 (OP Only) n=15,244 | 77,054 | 0 | 0 | 77,054 |
| 2 (OP & IP) n=2,750 | 34,653 | 14,650 | 0 | 49,303 |
| 3 (OP & ED) n=1,658 | 14,379 | 0 | 3,269 | 17,648 |
| 4 (OP & IP & ED) n=4,199 | 79,094 | 21,981* | 17,512* | 118,587 |
| **Total n=23,862** | **205,180** | **36,631** | **20,781** | **262,592** |

\*The 5,520 visits that encompass an ED attendance and an inpatient stay are included in both these cells.

In table 4 we show the median number of records made per visit day. We see that sub-cohort 4 (OP&IP&ED) again receives the largest number of records in all 5 categories. Patients in this sub-cohort receive a median of 29.90 measurement records per visit-day. In contrast patients in the OP only sub-cohort on average only receive records in the observations table at a median rate of 0.27 per visit-day. This difference may occur because inpatients are monitored throughout a stay with the recurrent capture of physiological measures such as blood pressure. The disparity becomes clear when we examine Figure 1. It is also worth noting that, despite also including IP stays, sub-cohort 3 (OP&IP) only have a median of 5.68 records per visit-day in contrast to 29.90 in sub-cohort 4 (OP&IP&ED). This may be accounted for by the fact that the median length of an inpatient stay for patients in sub-cohort 3 is 1 day, whereas in sub-cohort 4 it is 3 days. If a patient has an inpatient stay of only one day, for example as a day case for dialysis or chemotherapy, they are less likely to generate a large number of measurements during their stay.

**Table 4:** Median number of records made per visit-day.

| Sub-cohort | Condition | Drug | Measurement | Observation | Procedure |
| --- | --- | --- | --- | --- | --- |
| 1 (OP Only) n=15,244 | 0.00 | 0.00 | 0.00 | 0.27 | 0.00 |
| 2 (OP & ED) n=1,658 | 0.20 | 0.00 | 6.68 | 1.00 | 0.33 |
| 3 (OP & IP) n=2,750 | 0.53 | 0.00 | 5.68 | 1.00 | 0.36 |
| 4 (OP & IP & ED) n=4199 | 0.75 | 1.60 | 29.90 | 5.03 | 0.47 |

**Figure 1.**
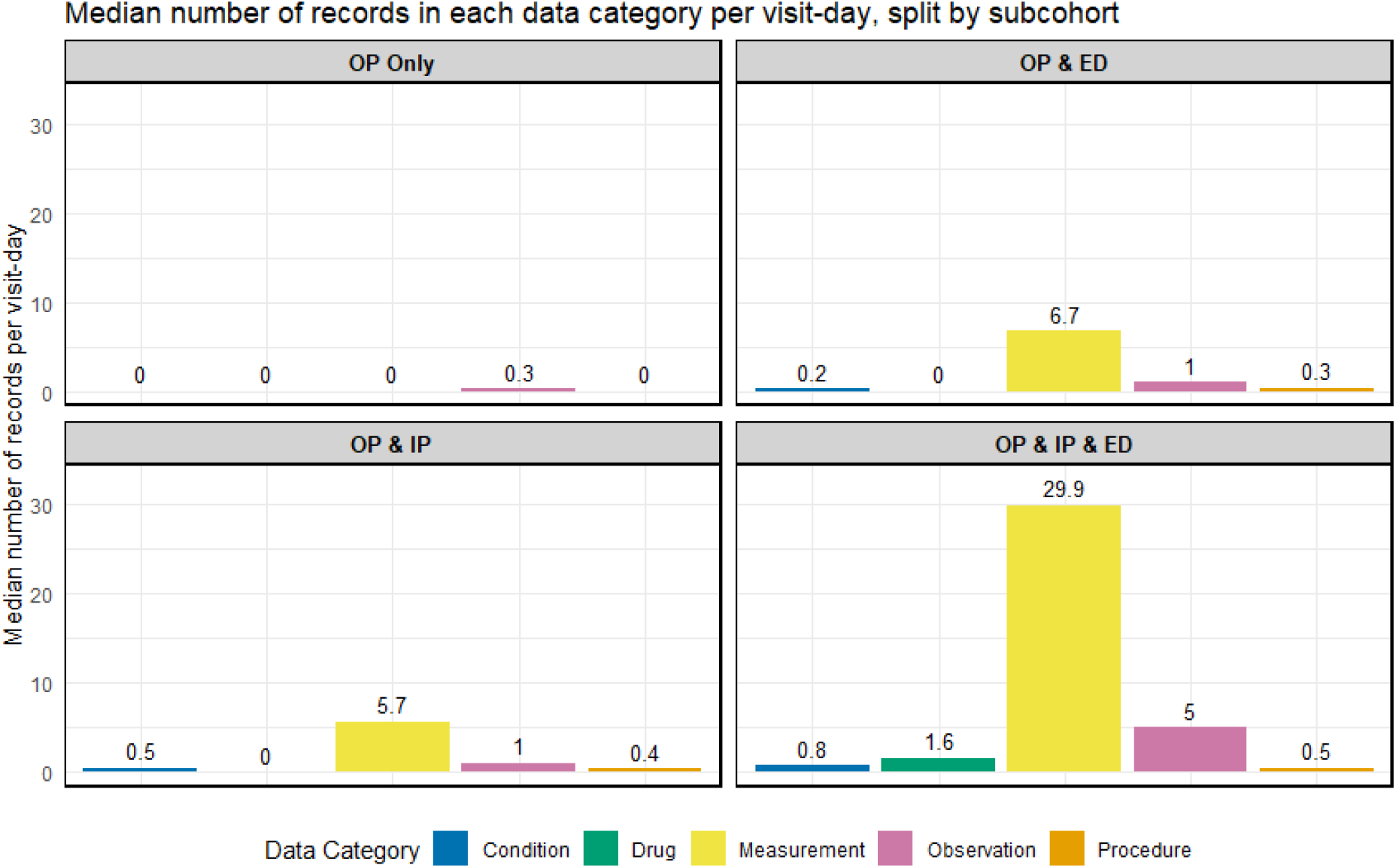
bar chart showing the median number of records made per visit-day in each of the data categories, per sub-cohort.

Table 5 shows for each data category the percentage of patients who have at least one record of that type. This is visualised as a waffle chart in figure 2. This clearly shows that the OP Only sub-cohort are almost entirely lacking any records for conditions and drugs, meaning that a clinician will not know from the currently OMOP-mapped medical records if a patient in this sub-cohort has a prior diagnosis or prescription. In contrast 99.69% patients in sub-cohort 4 (OP&IP&ED) have at least one condition recorded, and 78.66% have at least one record of a prescription. The percentages for conditions, measurements, observations and procedures are also high for sub-cohorts 2 and 3.

**Table 5:** For each data category, the percentage of patients who have at least one record of that type.

| Sub-cohort | Condition | Drug | Measurement | Observation | Procedure |
| --- | --- | --- | --- | --- | --- |
| 1 (OP Only) | 0.02 | 0.06 | 20.24 | 55.02 | 24.68 |
| 2 (OP & ED) | 92.82 | 11.94 | 85.83 | 100.00 | 83.72 |
| 3 (OP & IP) | 94.18 | 31.64 | 80.55 | 91.38 | 94.76 |
| 4 (OP & IP & ED) | 99.69 | 78.66 | 99.33 | 100.00 | 99.43 |

**Figure 2.**
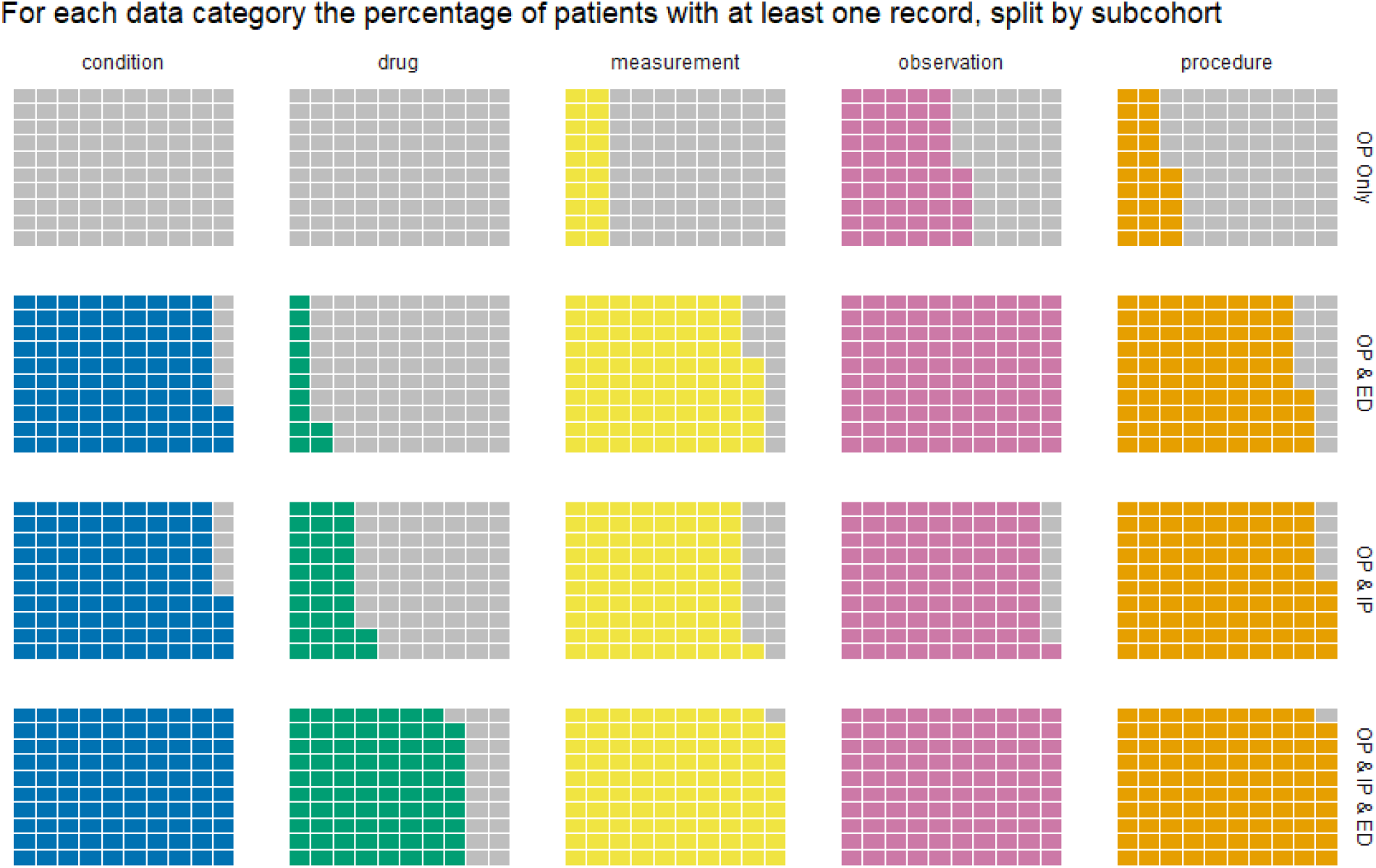
A waffle chart showing for each data category, the percentage of patients who have at least one record of that type.

## DISCUSSION

We have undertaken an investigation of the practicality of using the OMOP CDM to analyse data on neurology patients. We have quantified the volume and type of data available for neurology outpatient appointments and compared this to other visit types, and this has helped us to understand what type of data is currently available. This is important because it provides insight into the potential utility of the OMOP CDM database for a range of applications such as research, clinical, administrative and service planning functions, including the potential for large scale federated analytics.

The results show a stark contrast between the large volume of data collected for patients with inpatient stays or ED visits, and the paucity of data for patients who only attend the hospital for outpatient appointments. This is in part due to the fact that patients who attend only outpatients have fewer individual attendances than those who also visit as inpatients and/or attend ED. However, patients who attend only outpatient appointments also have far fewer records made per visit-day and have almost zero records regarding conditions or drugs. This leads to a large imbalance in the information available on a patient which depends on what type of appointments they have previously attended.

The virtual absence of information in this database on conditions or drugs for outpatients is of great concern. Knowledge of previous diagnoses and comorbidities, past and current prescribed medications, are key pieces of information required by a clinician at an outpatient appointment. Currently this information needs to be gleaned from the patient at the appointment, as it is not available in the database. This reduplicates information requests from patients and further pressurises scarce clinician time.

Furthermore, bias is introduced by virtue of the fact that when extracting patients with a particular diagnosis from the healthcare records for research purposes, only those who have required an ED visit or IP admission will have the diagnostic code in their record.

The patient population will therefore include those who have more severe neurological disease or complex comorbidities that require frequent ED and IP visits, while patients with milder neurological disease that is managed effectively in outpatient clinic will not be captured. This impedes our understanding of the true spectrum of disease severity for neurological disorders, and the identification of disease phenotypes that can help guide management and predict outcome.

There are a number of different reasons that some information is currently not available in the OMOP CDM database. Some data is available within the EPR or other hospital databases, but has not yet been transformed into the OMOP CDM, for example information regarding whether an outpatient appointment is new or follow-up. Other data is available in hospital databases, but isn’t yet supported by the OMOP CDM without using extensions to the current vocabulary, for example imaging data, although this should change with the addition of extensions to the OMOP CDM [16]. Yet other data is simply unavailable at the hospital, for example community prescribing data, although it is hoped that these data will be incorporated into the SDE in time.

The absence of diagnosis data presents a fundamental problem with respect to cohort definition. Much medical research relies on being able to identify patients by their diagnosis or condition and whether they have been prescribed a particular medication. The current lack of information in both of these categories for outpatients presents a barrier to conducting research on this population using the current OMOP CDM. This was a constraint in the present study as we were unable to stratify the sub-cohorts by condition, and therefore the 4 sub-cohorts were very heterogeneous. The availability of outpatient diagnosis data would facilitate future research and enable much more detailed analyses than we have been able to perform here.

During the analysis we found preliminary indications that inpatient length of stay affects the number of records per day. We noted a difference between the median number of records in sub-cohorts 3 and 4, both of which include inpatient stays. A possible explanation for this is that sub-cohort 3 includes more clinically stable elective patients, admitted directly onto an inpatient ward, whereas sub-cohort 4 includes patients admitted acutely unwell through the ED. Acutely unwell patients require more intensive monitoring by comparison with clinically stable patients, which may explain the higher median number of drugs, observations and measurements for this sub-cohort. Sub-cohort 3 also had substantially more inpatient stays of only one day compared with sub-cohort 4; again, this may be because sub-cohort 3 includes patients admitted electively for day case procedures, while sub-cohort 4 includes acutely unwell patients admitted through ED who are likely to need a longer stay. Future research should investigate this disparity further to determine whether admission acuity and/or the length of stay are driving this difference.

This study was limited by the absence of outpatient diagnostic coding which led to heterogenous sub-categories, as mentioned above. This lack of coding also led to the necessity of identifying the study population by the clinician recorded as conducting the initial outpatient appointment. This method of identifying neurology outpatients relies on the clinician being accurately recorded and of course will include patients for whom a non-neurological diagnosis was made at the appointment. If diagnoses from neurology outpatient appointments were to be coded, this would allow for a much more accurate identification of a study cohort. This study is also limited by the fact that this is a single centre study, however as many practices are the same across the NHS the results are unlikely to differ. Future research could be conducted once the OMOP CDM has been introduced to other regions to determine if the patterns we have observed here are replicated in other regions where mandated coding has not been introduced.

We have highlighted the large difference in the number of records created between different types of hospital visit, and also provided opportunity to reflect on the necessity of the amount and frequency of data capture in some hospital settings. Previous research has brought into question the efficacy of capturing large amounts of routine data [17,18], and questioned the necessity of ordering repeat tests [19,20]. These studies highlight the need to consider carefully the way in which records are made, the underlying activity that they represent, and the potential to save resources and improve patient care by eliminating unnecessary testing and data capture. Our findings highlight the difference in volumes of data recorded in outpatient vs inpatient or ED settings, but further research is needed to confirm how much of this data capture is necessary and whether systems of data capture could be made more efficient.

## CONCLUSION

Through this study we have shown that the OMOP CDM can be used to analyse the number and type of records captured for patients with different visit types. We found that outpatients have far less data recorded than inpatients or patients attending ED.

This highlights the need to improve data capture for outpatients, especially with regards to conditions and drugs as this is vital information both for the clinician when seeing a patient, and also for research. The lack of data available on outpatients risks limiting the opportunities for using data both to inform clinical services and research related to neurological conditions in an outpatient setting.

## Data Availability

Data used in the present work are not available for public sharing due to patient confidentiality. Access to the data may be considered under appropriate data-sharing agreements and ethical approvals.

## Notes

### Competing Interest Statement

The authors have declared no competing interest.

### Funding Statement

Fran Biggin is funded by The Engineering and Physical Sciences Research Council Impact Acceleration Account (EP/X525583/1). For the purpose of Open Access, the author has applied a CC BY public copyright license to any Author Accepted Manuscript version arising from this submission. The funders were not involved in study design, data collection, interpretation or writing of the report.

### Author Declarations

The Research and Innovation department of Lancashire Teaching Hospitals NHS Foundation trust gave approval for this work under a Service Evaluation agreement SE-475. The Service Evaluation process at LTHTR includes an evaluation of privacy protection and provides a conclusion as to whether a study requires further ethical review. The text of the registration agreement reads: "Service Evaluation is defined as being conducted solely to define or judge current care and does not require ethical review".

